# Temporal trends of stroke severity and the association between antithrombotic drug regimen changes and clinical outcomes after stroke in atrial fibrillation

**DOI:** 10.1101/2023.09.11.23295390

**Authors:** Jo-Nan Liao, Yi-Hsin Chan, Ling Kuo, Chuan-Tsai Tsai, Chih-Min Liu, Tzeng-Ji Chen, Gregory Y. H. Lip, Shih-Ann Chen, Tze-Fan Chao

## Abstract

**Background:** Stroke prevention is central to the management of patients with atrial fibrillation (AF) but the impact of NOACs on stroke severity from a nationwide perspective, and the impact of changes in antithrombotic regimen following an ischaemic stroke on subsequent clinical events is unceratin. The aims of the present study were as follows: (i) to describe the temporal trends in OAC use between 2012-2018, and the relationship to stroke severity at presentation; (ii) to describe antithrombotic therapy prescribing patterns following an ischaemic stroke, and the impact of post-stroke thromboprophylaxis on outcomes; and (iii) to assess the impact of changing OAC strategy in AF patients on a NOAC presenting with an ischaemic stroke.

**Methods:** From 2007 to 2018, a total of 63,365 patients were identified from the “National Health Insurance Research Database (NHIRD)” in Taiwan. The stroke prevention strategy before and after ischemic stroke and its association to stroke severity was analysed. Subsequent clinical events after ischaemic stroke included recurrent ischaemic stroke, intracranial haemorrhage (ICH), major bleeding, all-cause mortality and composite outcomes.

**Results:** The temporal trend disclosed that the overall OAC prescription rate was rising, with warfarin used declining and NOACs use increasing, which was associated with a gradual decline of moderate-severe and severe strokes. The post-stroke antithrombotic strategy was variable. Compared to NOACs post-stroke, there was a significant increase in ischaemic stroke and mortality in non-anticoagulated (adjusted hazard ratios [aHRs] 1.804 and 3.441, respectively) and antiplatelet users (aHRs 1.785 and 1.483, respectively). Warfarin use post-stroke was associated with a significantly incresaed risk of major bleeding compared to NOACs (aHR 2.839). Non-anticoagulated and antiplatelet users were associated with higher risks of both composite outcomes compared to NOAC. Among 769 patients who received NOACs before stroke and continued NOAC post-stroke, there was a higher risk of ischaemic stroke and composite outcomes with no difference in major bleeding, mortality or ICH if patients were changed to a different NOAC post-stroke.

**Conclusions:** In this nationwide cohort study, increasing use of NOACs was associated with a decline of moderate-severe and severe strokes. Compared to NOACs, non-anticoagulation and antiplatelet use were associated with a significant increase in ischaemic stroke, mortality, and the composite outcome with no significant differences in bleeding events. There was no significant difference of ischemic stroke, mortaltiy, and ICH between post-stroke warfarin and NOAC use but warfarin was associated with a significantly increased risk of major bleeding. A change of NOAC types after ischemic stroke was associated with a two-fold higher risk of ischaemic stroke and the composite outcomes.

## Introduction

Stroke prevention with oral anticoagulants (OACs) is the cornerstone for the management of atrial fibrillation (AF).^1^ Although the safety and effectiveness of the non-vitamin K antagonists (NOACs) are evident compared to vitamin K antagonists (VKA, e.g. warfarin), some patients taking OAC still present with ischaemic stroke events. While the residual risk of stroke and other cardiovascular events is well recognised, efforts to address this continued residual risk are needed. The impact on stroke severity at presentation with an ischaemic stroke may also differ between warfarin and NOACs, with implications from a nationwide perspective.

From the stroke prevention perspective, those who are non-anticoagulated (taking no antithrombotic therapy or antiplatelets) should be started on OAC, ideally a NOAC. Nonetheless, if the patient is already taking warfarin when presenting with an ischaemic stroke, the changes in thromboprophylaxis may have implications for prognosis. Also, clinicians are also uncertain what should be done if an AF patient taking a NOAC presents with an ischaemic stroke, i.e., whether the same NOAC should be continued, or changed to a different NOAC or warfarin, or whether antiplatelet therapy should be added.

Given these uncertainties, the aims of the present study were as follows: (i) to describe the temporal trends in OAC use between 2012-2018, and the relationship to stroke severity at presentaion; (ii) to describe antithrombotic therapy prescribing patterns following presentation with an ischaemic stroke, and the impact of post-stroke thromboprophylaxis on clinical outcomes; and (iii) to assess the impact of changing OAC strategy in those AF patients on a NOAC presenting with an ischaemic stroke.

## Methods

This study used the “National Health Insurance Research Database (NHIRD)” released by the Taiwan National Health Research Institutes. The National Health Insurance (NHI) system is a mandatory universal health insurance program that offers comprehensive medical care coverage to all Taiwanese residents. NHIRD consists of detailed health care data from >23 million enrollees, representing >99% of Taiwan’s population. In this cohort dataset, the patients’ original identification numbers have been encrypted to protect their privacy, but the encrypting procedure was consistent, so that a linkage of the claims belonging to the same patient was feasible within the NHI database and can be followed continuously. The descriptions about Taiwan NHIRD have been reported in our previous studies.^2-10^This study has followed the STROBE guidelines and patient consent was waived by the institutional review board of Taipei Veterans General Hospital.

### Study cohort and study design

From January 1, 2007, to December 31, 2018, a total of 427,625 AF patients aged ≥ 20 years were identified from NHIRD. AF was diagnosed using the International Classification of Diseases (ICD), Ninth Revision, Clinical Modification (ICD-9-CM) codes (427.31). The diagnostic accuracy of AF in Taiwan NHIRD has been validated previously.^11^ Among the study population, 63,365 patients who experienced ischaemic stroke have constituted the study cohort. They were further categorized into different year groups based on the date when they experienced ischaemic stroke. In the first part of our study, we investigated the associations between the trends of prescription rates of OACs, defined as the prescriptions of OACs (warfarin or NOACs) within 30 days before ischaemic stroke, and the severity of stroke among the whole study population (year 2007-2018). In the second part, we analyzed the associations between stroke prevention strategies after ischaemic stroke and the subsequent clinical outcomes among patients identified between year 2012 to 2018 and who survived longer than 90 days after stroke. We only focused on study subjects after 2012 since NOACs were available in Taiwan since then. The flowchart of study design and the number of AF patients experiencing ischaemic stroke at each year is shown in **supplemental Figure 1**.

### Severity of ischaemic stroke

The severity of ischaemic stroke was determined using the estimated National Institutes of Health Stroke Scale (eNIHSS) derived from the stroke severity index (SSI) proposed and validated in prior studies.^12-14^ The calculation rule of SSI was in the following^12^:

SSI = +3.5083 (airway suctioning) +1.3642 (bacterial sensitivity test) +4.1770 (intensive care unit stay) +4.5809 (nasogastric intubation) +2.1448 (osmotherapy such as mannitol or glycerol) +1.6569 (urinary catheterization) -5.5761 (general ward stay) +9.6804 (constant)

Then the eNIHSS can be transformed from SSI with the following equation^14^: eNIHSS = 1.1722 x SSI – 0.7533

The severity of ischaemic stroke was categorized as: minor (eNIHSS 0-4), moderate (5-15), moderate to severe (15-20) and severe (>20) stroke.^15^

### Stroke prevention strategies before and after ischaemic stroke

Patients were defined as “non-OACs,” “NOACs” and “warfarin” groups based on the stroke prevention treatments they received before ischaemic stroke. After ischaemic stroke, the stroke prevention strategies were classified as 6 groups: “none”, “antiplatelet agents (Ap)”, “warfarin”, “NOACs”, “warfarin + AP” and “NOAC + AP”. We reported the percentages of these 6 kinds of post-stroke treatments in 3 groups with different stroke prevention strategies before stroke.

### Subsequent clinical outcomes after Ischaemic stroke

We investigated the risks of several subsequent clinical events after ischaemic stroke, including recurrent ischaemic stroke, intracranial haemorrhage (ICH), major bleeding and all-cause mortality. Besides, four composite clinical outcomes were also studied, including “ischaemic stroke, ICH and mortality,” “ischaemic stroke, ICH, major bleeding, and mortality”, “ischaemic stroke and ICH” and “ischaemic stroke and major bleeding”. The risks of these events of patients receiving different stroke prevention strategies were compared to those who received NOAC alone (reference group). For patients who received NOACs before stroke and continued NOACs after ischaemic stroke, the risks of clinical events were compared between patients who stayed on the same NOACs and shifted to different NOACs.

### Statistical analysis

Data are presented as the mean value (standard deviation [SD]) for continuous variables and proportions for categorical variables. The differences between normally distributed continuous values were assessed using an unpaired 2-tailed t test or one-way analysis of variance (ANOVA) when the comparisons of multiple groups were performed. The differences between nominal variables were compared by Chi-square test. The risks of clinical events were assessed using the Cox regression analysis adjusted for age, sex, CHA_2_DS_2_-VASc score, HAS-BLED score, eNIHSS score and clinical factors which were not included in the CHA_2_DS_2_-VASc and HAS-BLED scores with a p value <0.05 between comparison groups. All statistical significances were set at a *p* < 0.05.

## Results

### Clinical characteristics

Clinical chararacteristics are summarised in Table 1, and the study flow in supplemental Figure 1. Most patients were aged ≥75 (66.9%) and at high stroke and bleeding risks based on the CHA_2_DS_2_-VASc and HAS-BLED scores, respectively. The mean eNIHSS stroke score was 12.79 (7.66).

**Table 1.** Clinical characteristics of patients when they experienced ischaemic stroke.

| Variables | n = 63,365 |
| --- | --- |
| Age, mean (SD) | 77.59 (10.49) |
| Age ≥ 75 years, n (%) | 42400 (66.91) |
| Age 65-74 years, n (%) | 13324 (21.03) |
| Sex (male), n (%) | 32870 (51.87) |
| Comorbidities, n (%) |  |
| Hypertension | 55576 (87.71) |
| Diabetes mellitus | 26513 (41.84) |
| Heart failure | 32977 (52.04) |
| Prior stroke/TIA | 63365 (100) |
| Vascular diseases | 8604 (13.58) |
| COPD | 18859 (29.76) |
| Hyperlipidemia | 25692 (40.55) |
| Autoimmune diseases | 2855 (4.51) |
| Cancer | 7483 (11.81) |
| Abnormal renal function | 13348 (21.07) |
| Abnormal liver function | 9343 (14.74) |
| Anemia | 10437 (16.47) |
| History of bleeding | 20542 (32.42) |
| Alcohol excess/abuse, n (%) | 1007 (1.59) |
| CHA<sub>2</sub>DS<sub>2</sub>-VASc score, mean (SD) | 5.98 (1.37) |
| HAS-BLED score, mean (SD) | 3.9 (1.1) |
| eNIHSS stroke score, mean (SD) | 12.79 (7.66) |

AF = atrial fibrillation; COPD = chronic obstructive pulmonary disease; ICH = intracranial hemorrhage; NSAIDs = NSAIDs= non-steroidal anti-inflammatory drugs; OAC = oral anticoagulant; SD = standard deviation; TIA = transient ischaemic attack

### Temporal trends of OACs and stroke severity

The temporal trends of the prescription of OACs between 2007 and 2018 show that the overall OAC prescription rate was rising, with warfarin use declining 2011 onwards and increasing use of NOACs between 2012-2018. (Figure 1A) This was associated with a gradual decline in the prevalence of moderate-severe and severe strokes, and a higher rising proportion of mild strokes, that coincided with the increasing use of NOACs. (Figure 1B)

**Figure 1.**
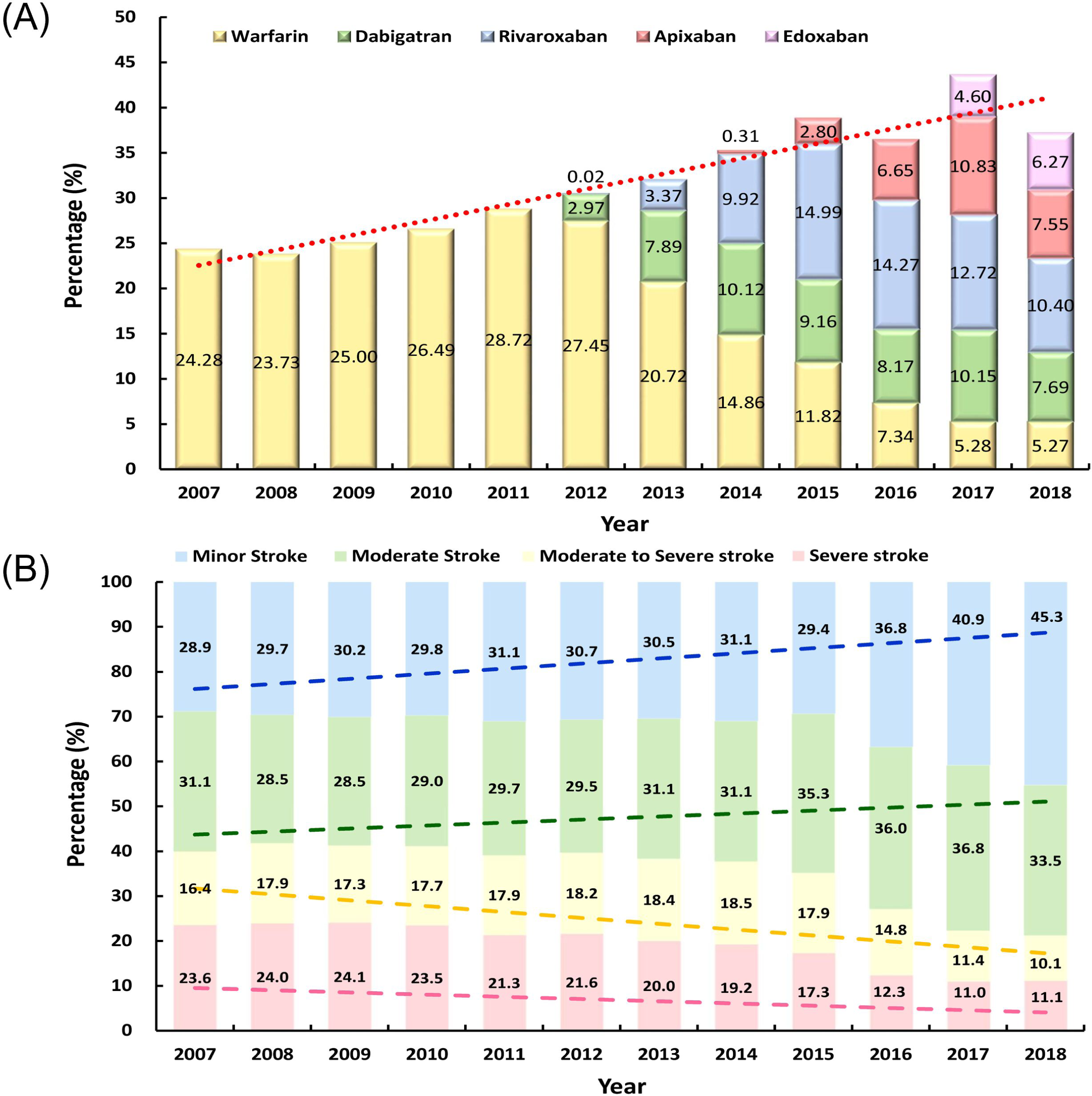
Temporal trends of the prescription rates of OACs before stroke and the severity of ischaemic stroke. (A) The overall OAC prescription rate was rising between 2007 and 2018, with warfarin use. declining 2011 onwards and increasing use of NOACs between 2012-2018. (B) A gradual decline in the prevalence of moderate-severe and severe strokes along with a higher rising proportion of mild strokes which coincided with the increasing use of NOACs was observed between 2007 and 2018. NOACs = non-vitamin K antagonist oral anticoagulants; OAC = oral anticoagulant

### Stroke prevention strategies before and stroke ischaemic stroke

Supplemental Table 1 shows the clinical characteristics in relation to the stroke prevention strategy after stroke between 2012-2018, where approximately 30% were either non-anticoagulated and nearly 20% taking NOACs (alone or in combination with antiplatelets) with the rest taking warfarin (alone or in combination with antiplatelets) or antiplatelets alone. The use of antithrombotic drugs after ischaemic stroke is summarized in Figure 2. In the non-anticoagulated subgroup at presentation, 33.51% were still non-anticoagulated and 39.19% were on antiplatelets post stroke. In those on NOAC when they had their stroke, most (42.72% and with antiplatelets, 13.78%) continued NOAC but 4.39% were changed to warfarin (alone or in combination with antiplatelets); 28.22% were non-anticoagulated. In those on warfarin at presentation, most remained on warfarin post stroke, with 12% changed to NOAC; 29.71% were non-anticoagulated.

**Figure 2.**
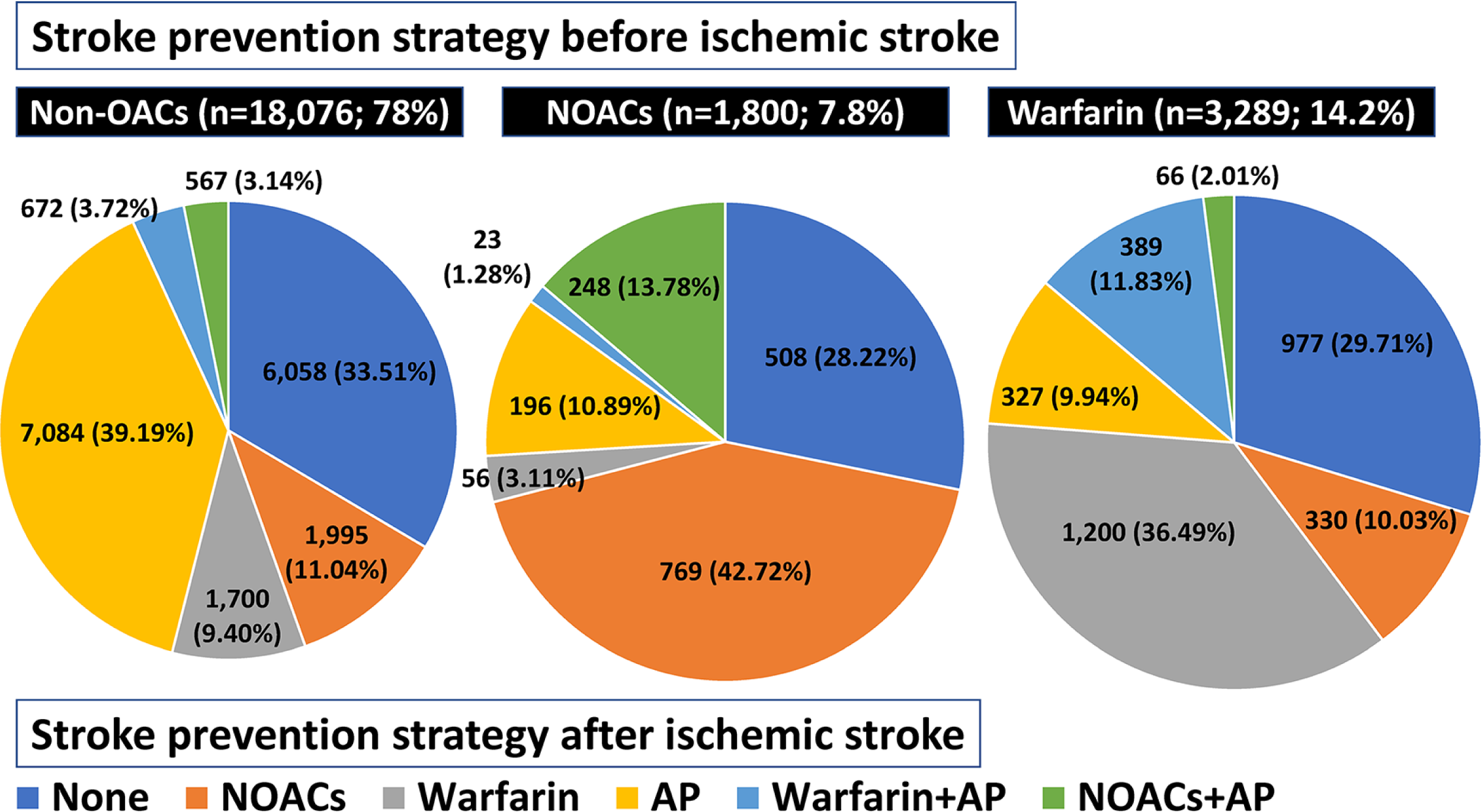
Stroke prevention strategies before and after ischaemic stroke. The use of antithrombotic drugs after ischaemic stroke were different among groups of different stroke prevention strategies before the index stroke event. AP = antiplatelet agents; NOACs = non-vitamin K antagonist oral anticoagulants; OAC = oral anticoagulant

### Risks of clinical events in different groups with different stroke prevention strategies after stroke

Figure 3 shows the impact on clinical outcomes in relation to post-stroke antithrombotic strategy. Compared to NOACs (reference), there was a significant increase in ischaemic stroke in non-anticoagulated and antiplatelet users (both adjusted hazard ratio [aHR] approximately 1.8), with no significant differences to warfarin. The same was evident for a higher mortality in non-anticoagulated and antiplatelet users (aHRs 3.441 and 1.483. respectively), with no significant differences to warfarin. Addition of antiplatelets to NOAC or warfarin resulted in no significant differences in ischaemic stroke or mortality.

**Figure 3.**
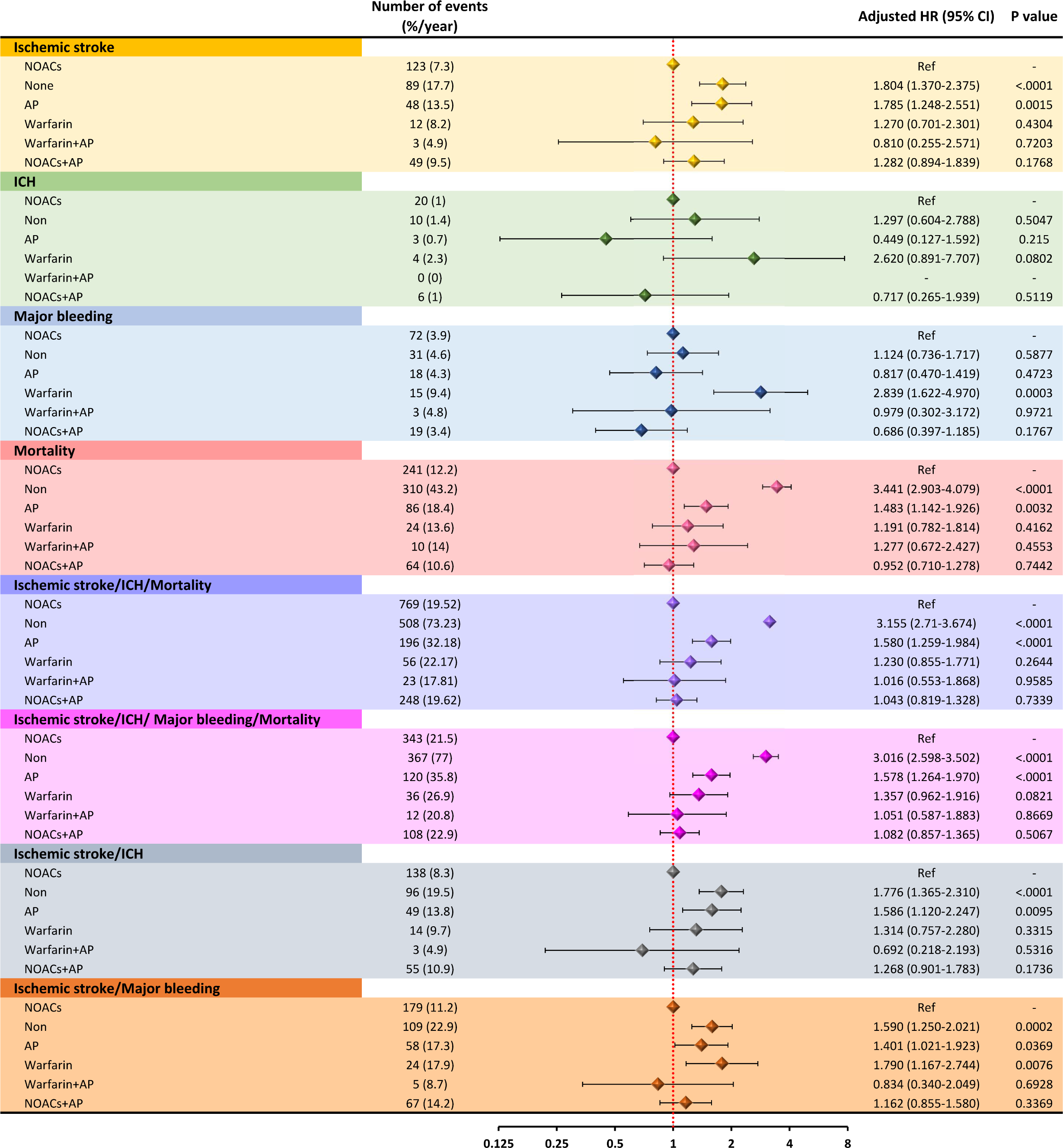
Risks of clinical events with different stroke prevention strategies after ischaemic stroke. AP = antiplatelet agents; CI = confidence interval; HR = hazard ratio; ICH = intra-cranial hemorrhage; NOACs = non-vitamin K antagonist oral anticoagulants

Compared to NOACs, there was a significant increase in major bleeding events post stroke for those on warfarin (aHR 2.839, 95% confidence interval [CI] 1.622-4.970), with no statistical difference in the other subgroups, including those non-anticoagulated. For ICH, when compared to NOACs, there was a trend for higher ICH with warfarin (aHR 2.620, 95% CI 0.891-0.707, p=0.0802) but no significant difference for other antithrombotic subgroups, including those non-anticoagulated.

We examined the impact on the composite outcomes: ‘ischaemic stroke, ICH and mortality’ and ‘ischaemic stroke, ICH, major bleeding and mortality’. When compared to NOAC (reference), non-anticoagulated and antiplatelet users were associated with higher risks of both composite outcomes, with aHR for non-anticoagulated and antiplatelet users being approximately 3.1 and 1.58, respectively. There was also trend for a higher aHR for ‘ischaemic stroke, ICH, major bleeding and mortality’ for warfarin users (aHR 1.357, p=0.0821).

Finally, we compared outcomes if NOAC users were continued on the same NOAC or changed to a different NOAC. Supplemental Table 2 shows the clinical characteristics of 769 patients who received NOACs before stroke and were continued on NOAC after stroke; most continued the same NOAC, but 144 patients were changed to a different NOAC. Those who were shifted to different NOACs had a higher eNIHSS score than those on the same NOACs. If patients were changed to a different NOAC post stroke, this was associated with a higher risk of ischaemic stroke (aHR 2.07) as well as the composite outcomes (aHRs between 1.359-1.849), with no difference in major bleeding, mortality or ICH (Figure 4).

**Figure 4.**
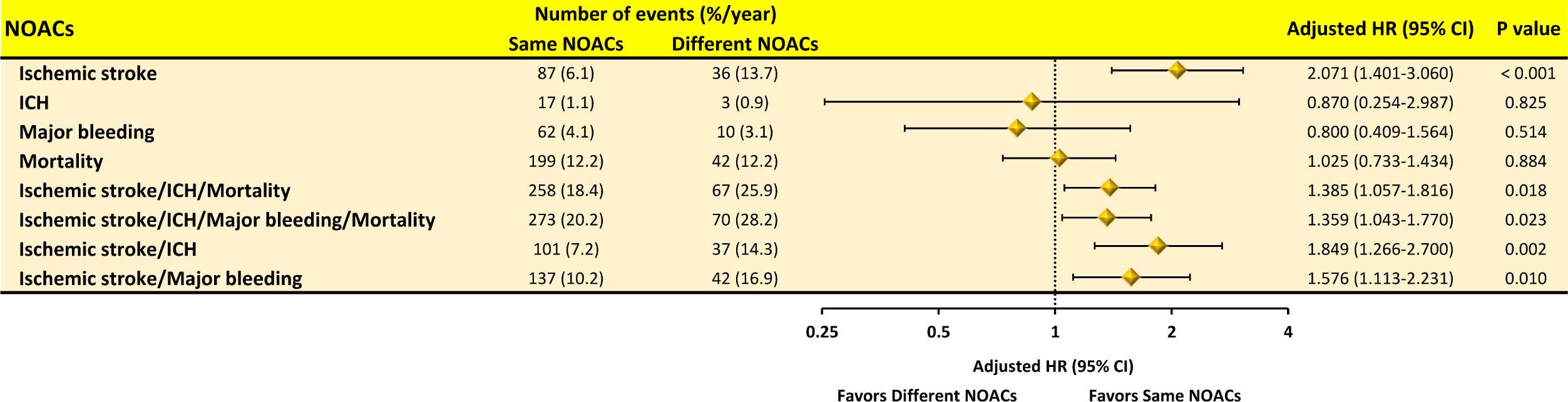
Risks of clinical events of patients who stayed on the same or shifted to different NOACs. We compared outcomes if NOAC users were continued on the same NOAC or changed to a different NOAC. A higher risk of ischaemic stroke as well as the composite outcomes was observed if patients were changed to a different NOAC post stroke, with no difference in major bleeding, mortality or ICH.

CI = confidence interval; HR = hazard ratio; ICH = intra-cranial hemorrhage; NOACs = non-vitamin K antagonist oral anticoagulants

## Discussion

In this paper, our principal findings are as follows: (i) Temporal trends of the prescription of OACs between 2007 and 2018 show that warfarin used declined 2011 onwards, with increasing use of NOACs between 2012-2018; these changes were associated with a gradual decline in the prevalence of moderate-severe and severe strokes, and a higher rising proportion of mild strokes, that coincided with the increasing use of NOACs; (ii) In the non-anticoagulated subgroup at presentation with ischaemic stroke, 33.51% were still non-anticoagulated and 39.19% were on antiplatelets post stroke; (iii) Compared to NOACs use post stroke, there was a significant increase in ischaemic stroke and mortality, as well as the composite outcome, in non-anticoagulated and antiplatelet users, with no significant differences to warfarin; (iv) Compared to NOACs, there was an increase in major bleeding and ICH events post stroke for those on warfarin, and (v) If NOAC patients were changed to a different NOAC post stroke, this was associated with a two-fold higher risk of ischaemic stroke as well as the composite outcomes.

The present study illustrates how the introduction of NOACs has changed the landscape for stroke prevention in AF, with the increase in OAC use overall, driven largely by more NOAC use.^16^ More importantly, it also shows a nationwide decline in moderate-severe and severe strokes over the same time period. Various smaller studies from other cohorts have reported that NOAC users presented with less severe stroke severity, when compared to warfarin or non-anticoagulated cohorts.^17-20^ Unfortunately, amongst the non-anticoagulated subgroup at presentation with ischaemic stroke, a third remained non-anticoagulated and nearly 40% were on antiplatelets post stroke. As shown in our study, this was associated in a substantial increase in risk of recurrent ischaemic stroke, as well as mortality, if OACs were withheld.

While OAC is often withheld due to a perceived risk of bleeding, we found no significant differences in major bleeding or ICH between NOACs and the non-anticoagulated subgroups. Nevertheless. the non-anticoagulated subgroup was at substantial risk of recurrent ischaemic stroke and mortality, with no significant difference in major bleeding or ICH compared to those started on NOACs post stroke. In the ELDERCARE-AF trial, there was no difference in major bleeding and ICH when low dose NOAC was compared to placebo.^21^ In elderly AF patients at high risk of bleeding, an observation we have previously reported from a nationwide cohort study also showed that NOACs were associated with a lower risk of ischaemic stroke, mortality, and the composite endpoint when compared with non-anticoagulated patients, while the risk of ICH and major bleeding were similar.^9^ Even if there is an increase in bleeding risk, as assessed by the HAS-BLED score, the use of NOAC conferred benefit in relation to clinical events, compared to OAC discontinuation.^7^

Unsurprisingly, there was an increase in major bleeding and ICH events post stroke for those on warfarin post stroke, when compared to NOAC treated patients. This is consistent with the large body of evidence from randomised trials as well as real world observational cohorts showing the better safety associated with NOAC use compared to warfarin [**35807073**], even in those perceived at highe risk of bleeding eg with anaemia or thrombocytopenia.^5, 22^ Nonetheless, bleeding risk assessment and mitigation is multifactorial, depending on non-modifiable and modifiable bleeding risk factors,^23^ but remains a topic of concern in East Asians taking antithrombotic therapy.^24^

Our study is in support of contemporary guideline recommendations that the default should be to offer stroke prevention with OAC (with NOACs being the preferred option) unless the patient is low risk.^1, 25^ Given the dynamic nature of bleeding risk, efforts to mitigate modifiable bleeding risk factors and schedule regular review and follow up should be undertaken.^23^ This was shown prospectively in the bleeding analysis from the mAFA-II trial, where the intervention clusters had less major bleeding and an increase in OAC use at one year, compared to usual care.^26^

Notwithstanding the small numbers, the present study also explores what If NOAC patients who present with an ischaemic stroke were changed to a different NOAC post stroke. Such a change was associated with a two-fold higher risk of ischaemic stroke as well as the composite outcomes. Thus, if an AF patient presents with an ischaemic stroke while on NOAC, the initial priorities should be to assess if the patient has been compliant with their drugs, and whether other comorbidities are present. Given the increasingly recognised residual risk of stroke despite OAC,^27^ much attention has been directed to a more holsitic or integrated care approach to AF management.^28^ Such an approach, based on the ABC (Atrial fibrillation Better Care) Pathway has been associated with improved clinical outcomes compared to non-adherent patients, leading to its recommendation in international guidelines.^1^

### Study limitations

There are several limitations of the present study. First, this is a retrospective study in which residual confounding factors potentially influencing the decision of OAC post stroke and outcomes might still exist despite our effort trying to adjust various factors. Second, the strategy of OAC post stroke was determined by the physicians in charge of the patients but not in a randomized fashion, which is a common limitation of retrospective studies. Third dosing information is not available in the present study, and so was the information of time in therapeutic range of warfarin pre-stroke and post-stroke. Lastly, there is lack of granularity for some risk factors, e.g. BP levels, creatinine clearance, etc., which may partly be responsible for the decreasing prevalence of moderate-severe and severe strokes over time.

## Conclusion

In this nationwide cohort study, increasing use of NOACs between 2012-2018 was associated with a decline in the prevalence of moderate-severe and severe strokes. Compared to NOACs, there was a significant increase in ischaemic stroke and mortality, as well as the composite outcome, in non-anticoagulated and antiplatelet users post stroke, with no significant differences in bleeding events. If NOAC patients presenting with a stroke were changed to a different NOAC post stroke, this was associated with a two-fold higher risk of ischaemic stroke as well as the composite outcomes.

## Data Availability

The data underlying this article are available in the article and in its online supplementary material.

## Acknowledgments

1. This work was supported in part by grants from the Ministry of Science and Technology (MOST 110-2314-B-075-059, MOST 111-2314-B-075-004-MY2) and Taipei Veterans General Hospital (V111C-020, V112C-019), Taipei, Taiwan.

2. This study is based in part on data from the National Health Insurance Research Database provided by the Bureau of National Health Insurance, Department of Health and managed by National Health Research Institutes. The interpretation and conclusions contained herein do not represent those of Bureau of National Health Insurance, Department of Health or National Health Research Institutes.

